# LSD increases sleep duration the night after microdosing

**DOI:** 10.1101/2023.06.27.23291970

**Authors:** Nathan Allen, Aron Jeremiah, Robin Murphy, Rachael Sumner, Anna Forsyth, Nicholas Hoeh, David B Menkes, William Evans, Suresh Muthukumaraswamy, Frederick Sundram, Partha Roop

## Abstract

Microdosing psychedelic drugs, at a level below the threshold to induce hallucinations, is an increasingly common lifestyle practise. However, the effects of microdosing on sleep have not been previously reported. Here we report results from a Phase 1 randomised controlled trial in which 80 healthy adult male volunteers received a six week course of either LSD (10 µg) or placebo with doses self-administered every third day. Participants used a commercially available sleep/activity tracker for the duration of the trial. Data from 3231 nights of sleep showed that on the night after microdosing participants in the LSD group slept an extra 24.3 minutes per night (95% Confidence Interval 10.3 - 38.3 minutes) compared to placebo - with no reductions of sleep observed on the dosing day itself. There were no changes in the proportion of time spent in various sleep stages or in participant physical activity. These results show clear modification of the physiological sleep requirements in healthy volunteers who microdose, and may have implications for the proposed therapeutic effects of microdosing in mood disorders such as major depressive disorder where sleep is frequently disturbed. The clear, clinically significant, changes in objective measurements of sleep observed are difficult to explain as a placebo effect.

## Introduction

The relatively recent emergence of the practice in which users self-administer psychedelic drugs, mainly lysergic acid diethylamide (LSD) or psilocybin, in repeated doses at levels below the threshold for overtly causing hallucinations is termed microdosing (Fadiman, 2011). Microdosers claim that this practice can improve mood and well-being, reduce the symptoms of both anxiety and depression, as well as potentially enhancing creativity and productivity (Anderson, Petranker, Christopher, et al., 2019; Anderson, Petranker, Rosenbaum, et al., 2019; Hutten et al., 2019; Lea et al., 2020; Rootman et al., 2021). While the veracity of many of these claims has yet to be adequately tested in randomised controlled trials in clinical populations (Polito & Liknaitzky, 2022), it is becoming increasingly clear that microdosing can cause distinct changes in neurophysiological function with reported changes in both resting and task-based EEG (Federico Cavanna et al., 2022; Glazer et al., 2023; Murray et al., 2022) and fMRI-based connectivity (Bershad et al., 2020) but there are no reports of changes in objectively measured behaviours. The potential effects of microdosing on sleep behaviour has not yet been thoroughly investigated. In fact, the effects of even macrodoses of psychedelic drugs on sleep function has received only sporadic investigation in the literature over the years. In a study of the N,N-dimethyltryptamine (DMT) containing brew ayahuasca (Barbanoj et al., 2008), healthy male participants (n=22) were administered placebo, ayahuasca (1 mg DMT /kg) or d-amphetamine in the daytime in a crossover design with polysomnography recorded the night of administration. Ayahuasca did not subjectively impair sleep quality but decreased the amount and relative proportion of time spent in REM sleep, increased REM onset latency and enhanced power of slow-wave NREM sleep. Similarly, in another crossover design study (Dudysova et al., 2020), participants (n=20) were administered either psilocybin (0.26 mg/kg) or placebo in the morning with polysomnography recorded in the evening. Results showed that psilocybin increased REM onset latency with a trend to decreased REM duration while sleep latency, total sleep time and the number of sleep cycles were not affected. These human studies concur with animals studies of the serotonergic psychedelics LSD, 2,5-Dimethoxy-4-iodoamphetamine (DOI) and mescaline, which have been found to generally increase wakefulness and decrease both REM and NREM durations (Colasanti & Khazan, 1975; Kay & Martin, 1978; Monti & Jantos, 2006).

With respect to microdosing, there is only a single report of objective sleep measurements available. Published in 1966, Muzio et al. (Muzio et al., 1966) administered participants low doses of LSD (range 4-40 µg) just prior to, or one hour after participants went to sleep by briefly awakening them to administer the drug. Generally, LSD was found to significantly increase REM duration, with REM bursts interrupting slow wave sleep as well as increased body movements and arousal periods occurring during some REM sleep periods. However, while the doses delivered in this study do overlap with the modern microdosing range, the timing of drug administration relative to sleep is quite inconsistent with modern microdosing practises which generally involve daytime microdosing.

Several microdosing studies have provided some subjective reporting around sleep quality. These studies tend to show bidirectional effects, with both improvements and difficulties in sleeping reported. Interviews with a small sample (n=24) of community microdosers found that overdosing and insomnia are common challenges of microdosing (Johnstad, 2018). Similarly, in an online survey of 525 participants who were currently microdosing 45% reported having had trouble sleeping ever with 3.2% saying it occurred often (Lea et al., 2020). On the other hand, a large cross-sectional study of community microdosers (n=3933) endorsed improving sleep as a motivation for microdosing (Rootman et al., 2021). Consistent with this, in an online cross-sectional survey (n=278) of microdosers, 28% of respondents found that microdosing improved their sleep quality (Anderson, Petranker, Christopher, et al., 2019). In a crossover-study of participants using psilocybin-containing truffles modifications to sleep patterns were noted by a small subset of participants (Marschall et al., 2022).

Overall, given the paucity of information regarding the effects of microdosing on objective sleep measures, in our recent Phase 1 microdosing trial (Murphy et al., 2023) using home-administration of LSD microdoses (10 µg base) in ecologically valid settings, all participants were given commercially available watches to wear to enable monitoring of sleep and activity patterns. The sensors in consumer-grade devices have been shown to be useful in assessing sleep stages, sleep-wake states and REM sleep, in naturalistic settings (Chinoy et al., 2022; de Zambotti et al., 2018). Participants microdosed at home every third day for six weeks with a one week baseline run-in period. Given that we planned to have 80 participants engaged in the protocol for approximately 49 nights, with fourteen doses per participant we had potentially ∼3920 sleep nights in the trial that could be analysed. Surprisingly, the results demonstrated that LSD microdosing caused robust and significant changes in REM sleep duration and total sleep duration the night after a microdose – with no effects observed on the night of the microdose.

## Methods

### Study Design

The design of the current (acronym: MDLSD) study was a double-blind parallel-groups trial. Healthy volunteers were randomised into LSD (n=40) and placebo (n=40) groups and self-administered 14 doses of either 10 µg LSD base (in water for injection) or inactive placebo (water only) by 1 mL oral syringe for sublingual administration every three days for six weeks. The study consisted of four visits for each participant: a screening visit at which they were assessed for eligibility, a baseline visit, followed by a first dosing visit (day 1) session seven days after the baseline visit and a final follow-up visit scheduled two days after the final microdose (day 42). Full protocol methods were prospectively published prior to any study visits occurring (Murphy et al., 2021) and a full description of the conduct of the trial has been previously published (Murphy et al., 2023). Only details relevant to the current results are recapitulated here. The trial was prospectively registered at ANZCTR (ACTRN12621000436875).

### Participants

80 healthy male participants were enrolled into the trial, with 40 participants randomised into the LSD group and 40 into the placebo group (see (Murphy et al., 2023) for CONSORT diagram). Key inclusion criteria included being male between the age of 25-60. Key exclusion criteria included: resting blood pressure over 160/90 mmHg, body weight < 50 kg or > 120 kg, significant renal or hepatic impairment, unstable medical or neurological conditions, lifetime history of depression/schizophrenia and psychotic disorders or current diagnosis of anxiety or eating disorders, suicidality, first degree relatives with a psychotic disorder, substance use disorder, use of psychotropic medication, use of a serotonergic psychedelic in the last year and any lifetime history of psychedelic microdosing. A urine drug test was performed at screening and a breathalyser test performed at the baseline and first dosing visits. Demographic characteristics of the population sample are shown in Table 1.

**Table 1:** Demographics of all randomised participants for both treatment groups in the MDLSD trial

| Observation | Placebo | LSD |
| --- | --- | --- |
| Age, *M (sd, range)* | 36.3 (7.2, 25-56) | 37.4 (9.2, 25-56) |
| Weight, kg, *M (sd, range)* | 83.3 (13.7, 55-112) | 85.1 (14.9, 64-129) |
| BMI, kg/m<sup>2</sup>, *M (sd, range)* | 25.9 (1.9, 19-34) | 26.4 (1.9, 20-39) |
| Lifetime serotonergic psychedelic use, Mdn (IQR) | 2 (0-7) | 3 (0-8.5) |
| Psychedelic naïve, *n (%)* | 12 (30) | 12 (30) |
| Ethnicity |  |  |
| Asian, *n (%)* | 4 (10.0) | 6 (15) |
| Latin American/Caribbean, *n (%)* | 3 (7.5) | - |
| Māori, *n (%)* | 2 (5) | 1 (2.5) |
| New Zealand European, *n (%)* | 29 (72.5) | 24 (60) |
| Other European, *n (%)* | 5 (12.5) | 11 (27.5) |
| Pasifika, *n (%)* | - | 4 (10) |
| Other, *n (%)* | 3 (7.5) | 1 (2.5) |

*Note:* All participants in this study are male. Ethnicity percentages will add up to more than 100 due to multiple ethnicities reported by single participants. More detailed demographic information is provided in Murphy et al. (Murphy et al., 2023).

### Procedures

Nearing the end of the baseline measurement session participants were given Fitbit Charge 3/4 devices and instructed to try to wear them for the remainder of the trial (apart from when they needed to charge the device or if it caused discomfort). The Fitbit app was installed on participant’s mobile devices to allow data synchronisation via Bluetooth. All notifications from the Fitbit app were turned off except for the low battery reminder. Typically the device required charging once per week. On dosing days participants were asked to dose before 11am to avoid any potential disruption to their sleep. Starting at 7am, participants received their first SMS reminder to take their microdose. They received hourly reminders until dose completion. For verifying dose compliance, participants were asked to video themselves taking the dose which they then uploaded to the study database. These videos were checked by the study team and then deleted (Murphy et al., 2023; Murphy et al., 2021). The video upload timestamps were used to establish the time of day each dose was taken.

### Data Analysis

The dataset for this study was initially available in JSON format and converted to text format using Python for easier manipulation and processing. Fitbit separates sleep into 4 states: “REM”, “Deep”, “Light”, and “Awake” where “Light” corresponds to polysomnography stages N1 + N2, “Deep” is stage N3, “REM” is REM sleep and “Awake” is Wake After Sleep Onset (WASO) (Dong et al., 2022). We computed variables “Asleep” as equal to (Deep + Light + REM) and Total as equal to (Deep + Light + REM + Awake) – the latter of which is similar to Total Sleep Time in polysomnography. The Fitbit sleep data was split across two files, presenting the potential for overlapping dates. 20 entries were identified with duplicate information, which were subsequently removed for accuracy. The sleep data, supplied by Fitbit, offered two methods to discern total sleep times: “Sleep Summary” and “Sleep Granular”. The Sleep Summary, gives the total time in minutes of each sleep state for each sleep. In contrast, the Sleep Granular data provides the duration and type of sleep transitions. An examination was performed of both methods to identify any discrepancies. The difference between the two sets was evaluated using the formula 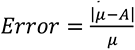, where 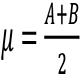 and *A* and *B* represent data from the summary and granular datasets respectively. The analysis revealed a mean error of 2.11 and a maximum error of 11.37 minutes for total REM sleep. Given the relatively small error magnitude, we opted to utilise the summary data for subsequent analysis but analysis of the granular data shows the same pattern of results presented here.

It is noteworthy that Fitbit assigns sleep data based on the sleep start date. A cursory assumption might suggest that the start sleep date corresponds accurately with the dosing days, but this was not always the case. To ensure accurate sleep date assignment, all sleep start times were plotted, as shown in Figure 1 to determine an appropriate “cut-off” time. The graph suggests that an appropriate cut-off time would be 9am, which roughly matches up with the expected microdosing time, where any sleep events before that would correspond to the previous night.

**Figure 1:**
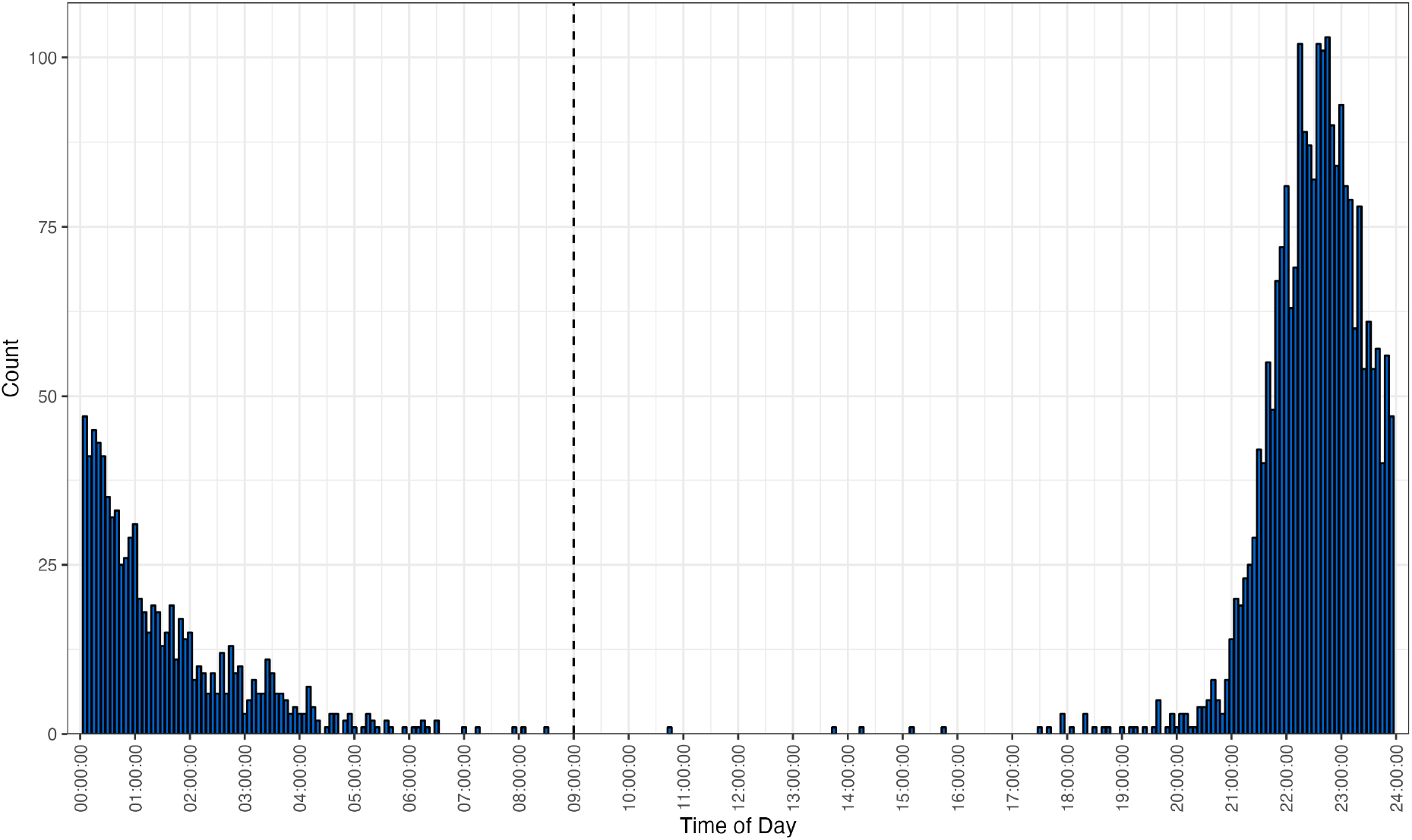
Distribution of participant sleep start times across the trial, grouped into 5-minute chunks. The cut-off time for a night’s sleep is indicated by the dashed line at 9:00am.

Finally, the sleep data provided by Fitbit consists of a variety of sleep times. The primary interest was in analysing the effects of LSD on sleep and to focus on how it affects a person’s main sleep, not any napping. As such, we employed Fitbit’s “isMainSleep” flag to filter out nap times. This flag is set to true if the sleep is the main sleep of the day and false if it is a nap. Notably, Fitbit may assign two main sleep periods if the main sleep is interrupted by a significant period of wakefulness, such as a trip to the toilet – these occurrences were merged.

### Statistical Methods

Statistical analyses were conducted using linear mixed-effects modelling using the lmerTest package in R (Kuznetsova et al., 2017) with Group (LSD, placebo) and Day (Dose, Dose + 1, Dose + 2) being treated as fixed effects with dummy coding (with the dosing day as the baseline condition), and participants as a random effect. The primary estimates of interest being the Group x Day interaction effect. All data from the trial was analysed using an intention-to-treat scheme with no imputation for missing data due to participant dropouts or missing sleep/activity data. There was a baseline period of seven nights of sleep, the average of which was used as a covariate in the analyses. Although the protocol was published prospectively, we had no pre-specified analyses or hypotheses relating to sleep and as such the analyses here should be considered exploratory. To correct for multiple comparisons, Bonferroni correction was applied to the alpha threshold for each analysis conducted depending on the number of comparisons conducted within an analysis.

## Results

Of the 80 participants who commenced the trial, five did not complete the study protocol. Four were discontinued from the trial due to adverse effects (mild anxiety) and one for reasons unrelated to the trial. A further three participants received an extra dose due to scheduling issues with follow-up and were in trial for an additional 2-4 nights. In Supplementary Figure 1 the total number of nights of sleep data are plotted for each participant. Overall there were 3231 nights of data available analysis including 503 nights of baseline data, 935 nights of dosing day data, 927 nights the day after dosing and 866 two nights after microdosing. The mean nights of data available per participant was 40.39 (sd=9.96).

Figure 2 presents the results of the linear mixed model analysis, with baseline adjustment, of time spent in each stage (Deep, Light, REM, Awake) as well as Asleep and Total. The overall pattern of effects showed relatively increased time for all metrics on the Dose+1 night but only time spent in REM sleep (p = 0.0037), Asleep (p = 0.0026) and Total (p = 0.0027) reached significance with the Bonferonni corrected significance threshold of 0.00833 (0.05/6). These differences corresponded to an extra 8.13 minutes of REM sleep (95% CI 3.34-12.9), 21.1 minutes Asleep (95% CI 8.9-33.2) and 24.3 minutes of Total sleep (95% CI 10.3-38.3) on the Dose+1 night for the LSD group, compared to the Placebo group. Similar results were observed when the average of the baseline run-in period was not included as a covariate in the analysis (see Supplementary Figure 2). Given this pattern of results, we then examined the proportion of time spent in each sleep stage – the results of which are displayed in Figure 3. These analyses showed that there were no changes which approached significance even with an uncorrected significance threshold of p=0.05.

**Figure 2:**
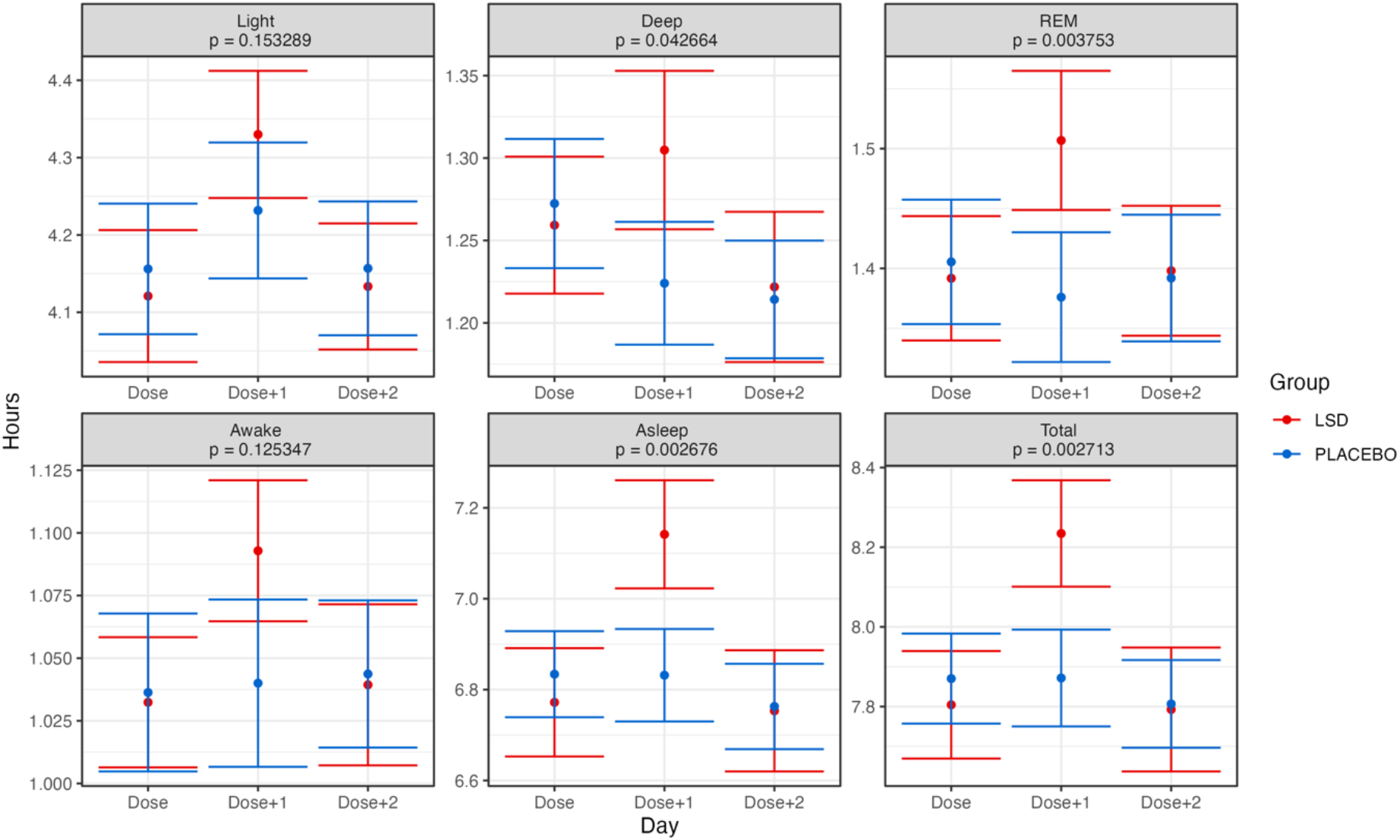
Grand average time spent in each of the sleep stages for each day (dose, dose+1 dose+2) and group in the trial. Error bars represent the standard error of the mean calculated across participants. Presented p values are for the Group x Day interaction effect. The Bonferroni-corrected alpha threshold for these analyses was 0.00833 (0.05/6). Note: Asleep = Deep + Light + REM. Total = Deep + Light + REM + Awake. See text for details.

**Figure 3:**
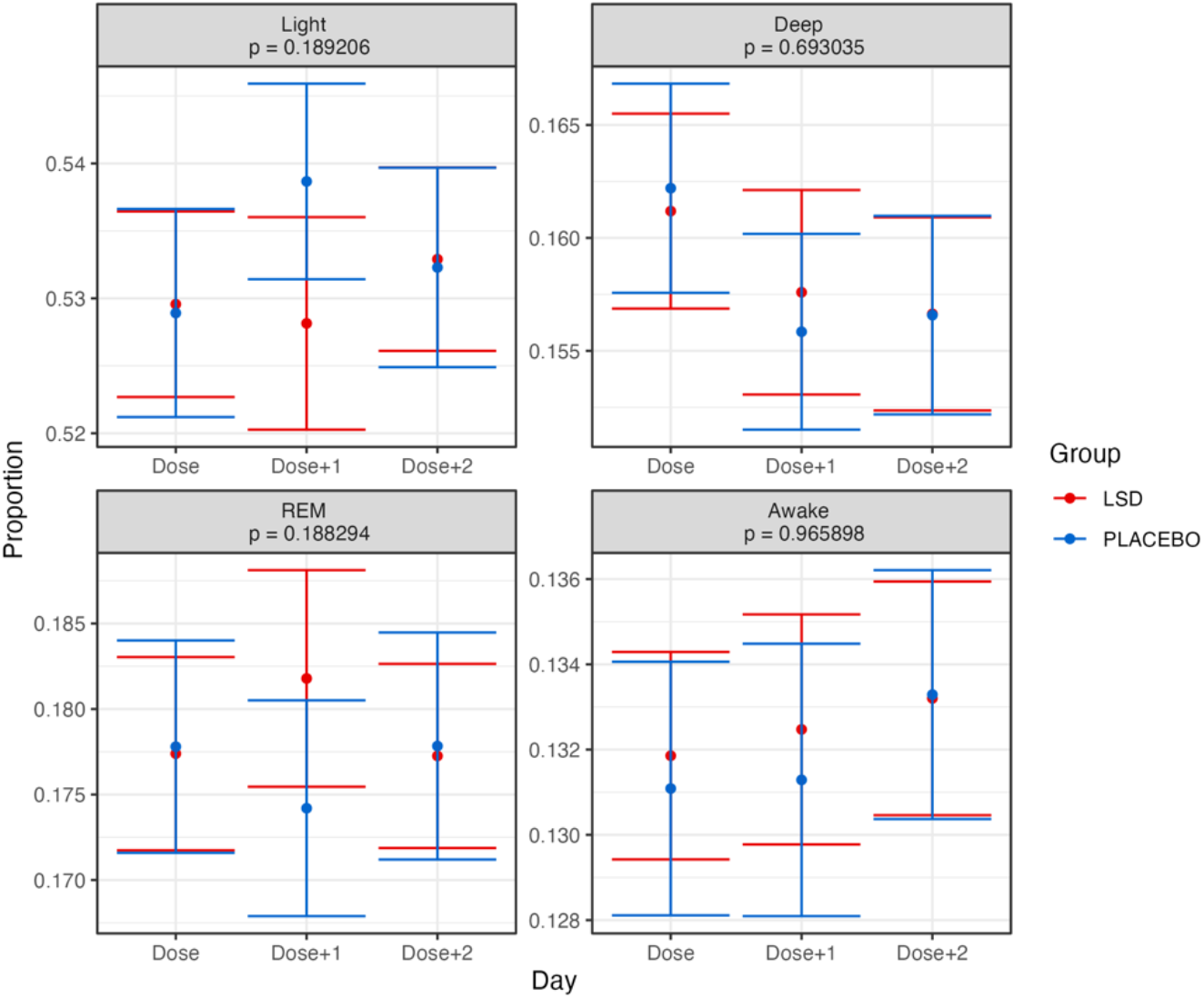
Grand average distribution of each sleep stage (as a proportion of total sleep) for each day (dose, dose+1 dose+2) and group in the trial. Error bars represent the standard error of the mean calculated across participants. Presented p values are for the Group x Day interaction effect. The Bonferroni-corrected alpha threshold for these analyses was 0.0125 (0.05/4). See text for details.

The observed pattern of sleep alterations was further explored by computing the time that participants went to sleep each night and the time that they woke up the next morning. These results (Figure 4) demonstrated that participants in the LSD group went to bed significantly (p=0.005) earlier on the Dose+1 night. This corresponded to going to bed an extra 25.17 minutes earlier on the Dose+1 night. No significant change was observed in the time at which participants woke up. It was also tested whether the observed modification in sleep time on the Dose+1 night changed across the time by adding dose number (1-14) to the mixed model. No effects across time were observed (see Supplementary Figure 3).

**Figure 4:**
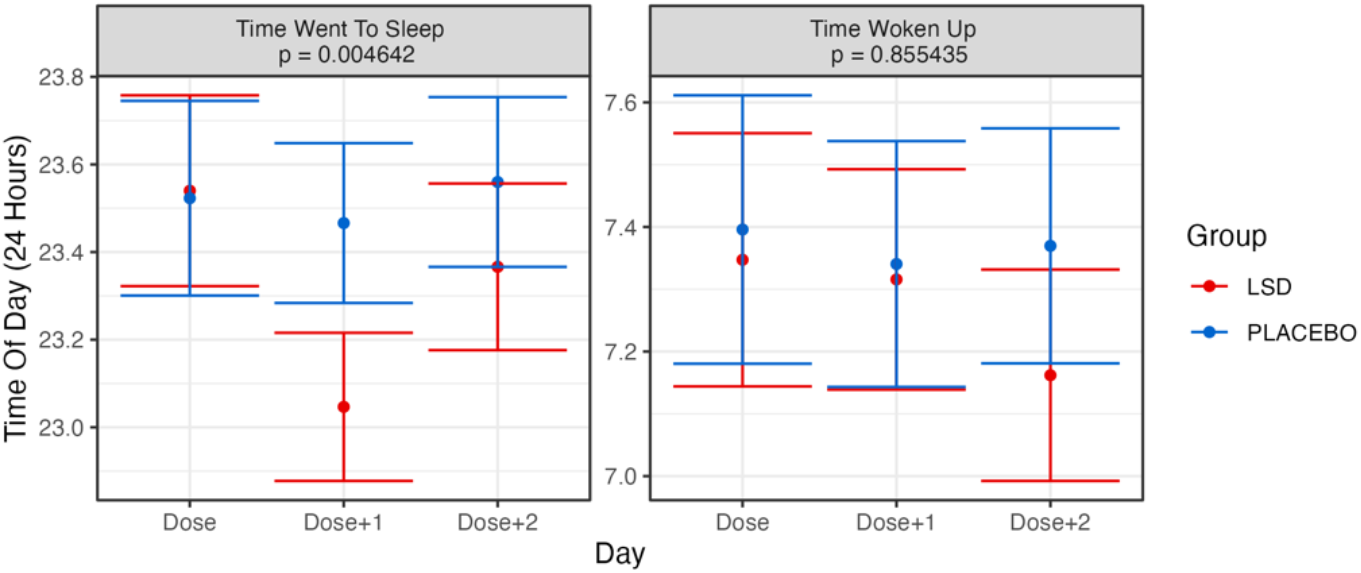
Grand averages for when participants went to sleep and woke up for each day (dose, dose+1 dose+2) and group in the trial. Error bars represent the standard error of the mean calculated across participants. Presented p values are for the Group x Day interaction effect. The Bonferroni-corrected alpha threshold for these analyses was 0.025 (0.05/2). See text for details

One possibility that might explain the extra Dose+1 sleep requirements was if the physical activity patterns of participants were modified by the drug. Similar to Supplementary Figure 1, Supplementary Figure 4 plots the total days of activity data available for analysis for each participant. There was slightly more activity data available for analysis than sleep data. Overall there were 3842 days of activity data available for analysis including 556 days of baseline data, 1102 days of dosing day data, 1101 days the day after dosing and 1083 two days after microdosing. The mean days of data available per participant was 45.67 (sd=6.62). In Figure 5, various activity pattern metrics (calories, distance, and steps) are plotted. These results show no noticeable or significant changes interaction effects. Furthermore, we also analysed the activity state provided from the wearable device in Figure 6 (sedentary, light activity, moderate activity, very active). As with the previous analysis, there were no significant interaction effects. Visually, participants in the LSD group appear to have overall lower moderately and very active minutes, however this was also visible in the group’s baseline averages and there was no main effect.

**Figure 5:**
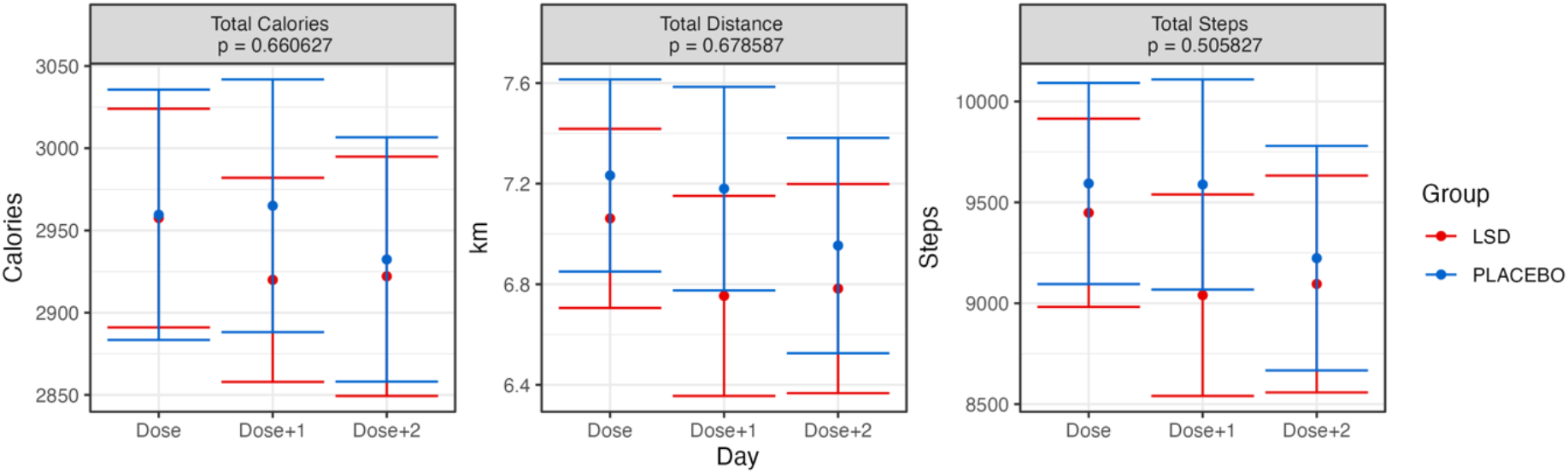
Grand average values for basic activity for each day (dose, dose + 1 dose + 2) and group in the trial. Error bars represent the standard error of the mean calculated across participants. Presented p values are for the Group x Day interaction effect. The Bonferroni-corrected alpha threshold for these analyses was 0.013 (0.05/3). See text for details.

**Figure 6:**
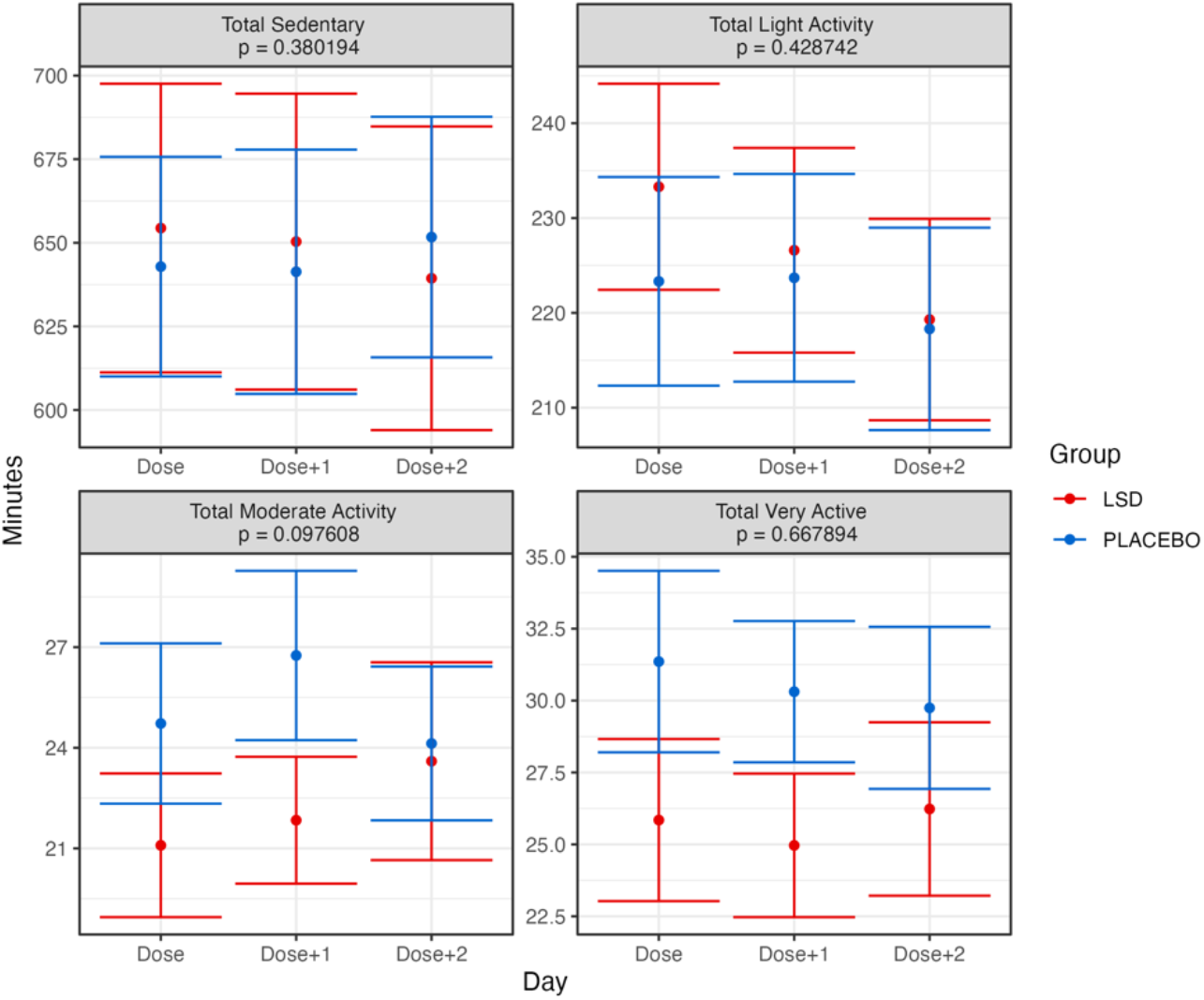
Grand averages of minutes spent in various states of activeness, as categorised by Fitbit, for each day (dose, dose + 1 dose + 2) and group in the trial. Error bars represent the standard error of the mean calculated across participants. Presented p values are for the Group x Day interaction effect. The Bonferroni-corrected alpha threshold for these analyses was 0.0125 (0.05/4). See text for details.

Although we did not include any subjective measurements of sleep quality, participants were asked to report their tiredness each night and were offered the chance to volunteer pertinent information in their daily report form and in a qualitative interview. Previously we reported a marginally significant effect (Murphy et al., 2023) where participants reported being more tired on Dose+1 days. Participants could also report sleep disturbances as an adverse event. 10 sleep-related adverse events were reported in the LSD group and 4 in the placebo group – which was not statistically significantly significant in terms of odds ratios (Murphy et al., 2023). Finally, at the end of the trial, participants took part in a semi-structured interview the full content of which we plan to publish elsewhere. Relevant to the current report, a number of participants in the LSD group commented that microdosing could both increase and decrease their energy levels (see Supplementary Materials for selected quotes). However, participants in the LSD group did not explicitly comment on needing extra sleep or going to bed earlier.

## Discussion

In this study it was found that participants in the LSD group had significantly increased sleep time compared to participants in the placebo group when they had taken a microdose the previous day, but no differences were found the night of the dose. These differences corresponded to an extra 8 minutes of REM sleep, 21 minutes of asleep time and 24 minutes of total sleep time the night after microdosing with no differences in sleep on the microdosing day itself with participants going to be earlier the night after microdosing. There were no differences in the ratio of time spent in each sleep stage, nor were there detectable differences in the physical activity of participants between the groups or evidence of tolerance/sensitisation.

The extra 24 minutes of sleep obtained by participants on the Dose + 1 night is not only a statistically significant difference but a clinically meaningful difference between the two groups, with 20 minutes of sleep speculated to be a clinically meaningful difference in terms of sleep duration (Singh et al., 2020). Practically speaking this result has implications for both the design of therapeutic microdosing protocols with LSD and their potential therapeutic mechanism of action. Pragmatically, the unexpected finding of the extra sleep required after microdosing suggests that it is important for microdosing protocols to have at least one day “off” between doses to ensure that patients are well rested/recovered before the next microdose is taken. While participants did report being marginally more tired on the day after dosing, in retrospect they did not explicitly mention requiring more sleep or going to bed earlier suggesting that these processes might have been occurring covertly. In line with reports from lifestyle users (Johnstad, 2018; Lea et al., 2020), some participants in this trial did retrospectively report some trouble sleeping on dose days, however inspection of adverse event data showed that difficulty sleeping was not reported at significantly higher rates than placebo and our objective data is consistent with this. This is likely because our home-administration dosing protocol required participants to have dosed by 11am to avoid any disruptions to sleep – a strategy that appears to have been largely successful in this study.

Anecdotally, microdosing users (Anderson, Petranker, Christopher, et al., 2019; Anderson, Petranker, Rosenbaum, et al., 2019; Hutten et al., 2019; Lea et al., 2020; Rootman et al., 2021) have consistently reported improvements in depressive symptomology. Modifications to sleep may be a factor that contributes to that effect. Difficulties with sleeping are commonly reported in mood disorders such as major depressive disorder (Plante, 2021; Staner, 2010) and premenstrual dysphoric disorder (Meers & Nowakowski, 2020). Polysomnography studies have shown that patients with depression show decreases in slow wave sleep, as well shortened REM onset latency and total REM sleep time (Riemann et al., 2020). It has generally thought that sleep difficulties and depression share a bidirectional causal relationship - although randomised controlled trials looking at sleep interventions to improve depression have only yielded mixed results (Plante, 2021). Synaptic plasticity theories of depression propose that depression is characterised by modifications of the circuitry underlying cortical adaptability. Sleep is a well-known modulator of synaptic plasticity (Diekelmann & Born, 2010) and it is thought that a critical function of sleep, particularly REM sleep, is to support the formation and consolidation of new memories (Peever & Fuller, 2017). In a large epidemiological study, patients with depression reported on average 40 minutes less sleep than non-depressed patients (Cepeda et al., 2016). Speculatively, a candidate therapeutic mechanism by which microdosing LSD might improve mood is by restoring sleep and promoting accompanying synaptic plasticity. In contrast to the current findings with microdosed LSD, most antidepressants actually suppress REM sleep and first-line treatments for depression such as selective serotonin reuptake inhibitors (SSRIS) can actually decrease sleep continuity leading to increased levels of insomnia (Riemann et al., 2020). As such, the potential use of microdosed LSD as an antidepressant may have very different effects on sleep, and therapeutic effects in patients with depression than standard antidepressants.

The current study adds to a growing body of knowledge of microdosing. Several studies using uncontrolled microdosing regimens have claimed that most of the anecdotal subjective effects of microdosing are probably related to participant expectancy and explainable as placebo effects (F. Cavanna et al., 2022; Federico Cavanna et al., 2022; Szigeti et al., 2021). Significant effects of microdosing on vital signs have been relatively inconsistent in controlled trials (Bershad et al., 2019; de Wit et al., 2022; Family et al., 2020; Hutten et al., 2020; Ramaekers et al., 2021), but central nervous system penetration of microdoses has been demonstrated multiple times using EEG (Federico Cavanna et al., 2022; Glazer et al., 2023; Murray et al., 2022) and fMRI (Bershad et al., 2020). To our knowledge this might be the first report of an effect of microdosing on an objectively measurable behavioural outcome and its completely unexpected nature is difficult to explain as a placebo/expectation effect, particularly given the null response in the placebo group data.

## Strengths and limitations

The use of a commercially available wearable device to track sleep and activity is both a strength and weakness of the current study. The major strength of using the fitness trackers in this context is the ability to collect a large amount of naturalistic sleep data (3231 nights worth) while being relatively unintrusive for participants compared to spending nights in a sleep laboratory. For example, and by contrast, the psilocybin sleep study of Dudysova et al. (Dudysova et al., 2020) included 40 nights of sleep data recorded the night of after a large dose of psilocybin was administered. At the same time, the wearable devices used are limited in that they do not provide reliable access to the important clinical metrics of sleep/REM onset latency. The other limitation of commercial wearable devices are the “black box” nature of the transfer functions, from their physiological sensors to the sleep metrics which they provide end-users for analysis. That said, a number of studies have directly compared Fitbit devices to polysomnography with a meta-analysis of eight studies (Haghayegh et al., 2019) concluding that they are accurate and reliable in terms of detecting sleep durations and staging although still inferior to polysomnography.

Other limitations of the current study as we have previously noted include the use of only male participants with relatively homogeneous ethnic representation and the fact that this cohort was healthy with no reported mental health or sleep issues. Finally, the current study did not include any subjective scales of sleep quality which would be useful to include in future microdosing studies.

## Conclusion

Given the significant modification in total sleep observed here with LSD microdosing and the potential clinical implications, this result provides a strong justification to incorporate wearable devices for sleep monitoring in our upcoming Phase 2 trials of LSD microdosing in patients with major depressive disorder (“https://www.anzctr.org.au/Trial/Registration/TrialReview.aspx?id=385758,”).

## Data Availability

Data produced in the present study are available upon submitting a research proposal to the authors and upon signing of confidentiality agreements with The University of Auckland

## Acknowledgements and Disclosures

The research was funded by donations from 3 individual donors and by a research grant from the Health Research Council of New Zealand (Grant No. 20/845 to SM and RS).

SM, RS, NH, NA, AJ, FS, and PR have received research funding from MindBio Therapeutics Ltd. to support data analysis.

SM has received funding from atai Life Sciences for unrelated research work.

No other authors report biomedical financial interests or conflicts of interest.

## Author Contributions

SM, RM, RS, DM, WE, NH and AF implemented and executed the study, with input from FS, and PR. NA and AJ performed all results generation and figure creation. SM, NA, and AJ wrote the manuscript.

## Trial Registration

Australian New Zealand Clinical Trials Registry: A randomised, double-blind, placebo-controlled trial of repeated microdoses of lysergic acid diethylamide (LSD) in healthy volunteers; https://www.anzctr.org.au/Trial/Registration/TrialReview.aspx?id=381476; ACTRN12621000436875.

## Participant Quotes

### Decreased energy

1. I do find on those dose days, that as I might have mentioned, I have lower energy, and my body feels a bit- like even, even when I mentally feel stimulated, my body felt a bit weak kind of throughout the day. So on those dose days, I was always a lot less motivated to do like a workout [Participant A, LSD]
2. Right. I think as far as energy level goes, like, definitely post-dosage I could definitely feel a low and I just felt more tired as if the dosing consumed a higher level of energy than normal. And so yeah, yeah just a little fatigue.[…] But particularly the next day afterwards. Yeah. But I think by the second or third day, I was normalising […] I definitely noticed extra fatigue for sure….For me, I would notice it the next morning, yeah. The same night it’s a bit hard to differentiate, but that’s just the evening fatigue. But the next morning is definitely, I can notice like, I slept like- I felt like I needed, like, to sleep in and stuff. [Participant B, LSD]
3. But never, it was probably- it was, I found it quite emotionally draining. […] I probably had less energy on the days that I took it actually. […]it was just probably just like, more of a lack of focus [Participant C, LSD]

### Increased energy

1. It gave me like a good energy I think. It made me want to do things and like yeah, just like get into stuff and just go and do all these things that I wanted to do. I found it yeah, definitely… it gives you an interesting clarity.[…] And took out any of the buzz that was in the background and you can just deal with stuff as it came. [Participant D, LSD]
2. Except from that, I did feel like the, in terms of like energy levels. I mean, I mean, physical energy, I did feel when increase that. So I would maybe prefer, in the dose days do some maybe physical activities […] because it felt, that I had that extra energy. [Participant E, LSD]
3. Yeah, so… I felt very, like it was interesting, because I had like, huge rushes of energy Or I found myself- and maybe it’s something to do with like the energy I felt- I found myself staying up later than I kind of wanted to. But equally, I didn’t feel overly unrested the next day. Like, I know, I normally needed more sleep than I actually got often. But yeah, I didn’t feel overly tired. [Participant F, LSD]

---

**Supplementary Figure 1:**
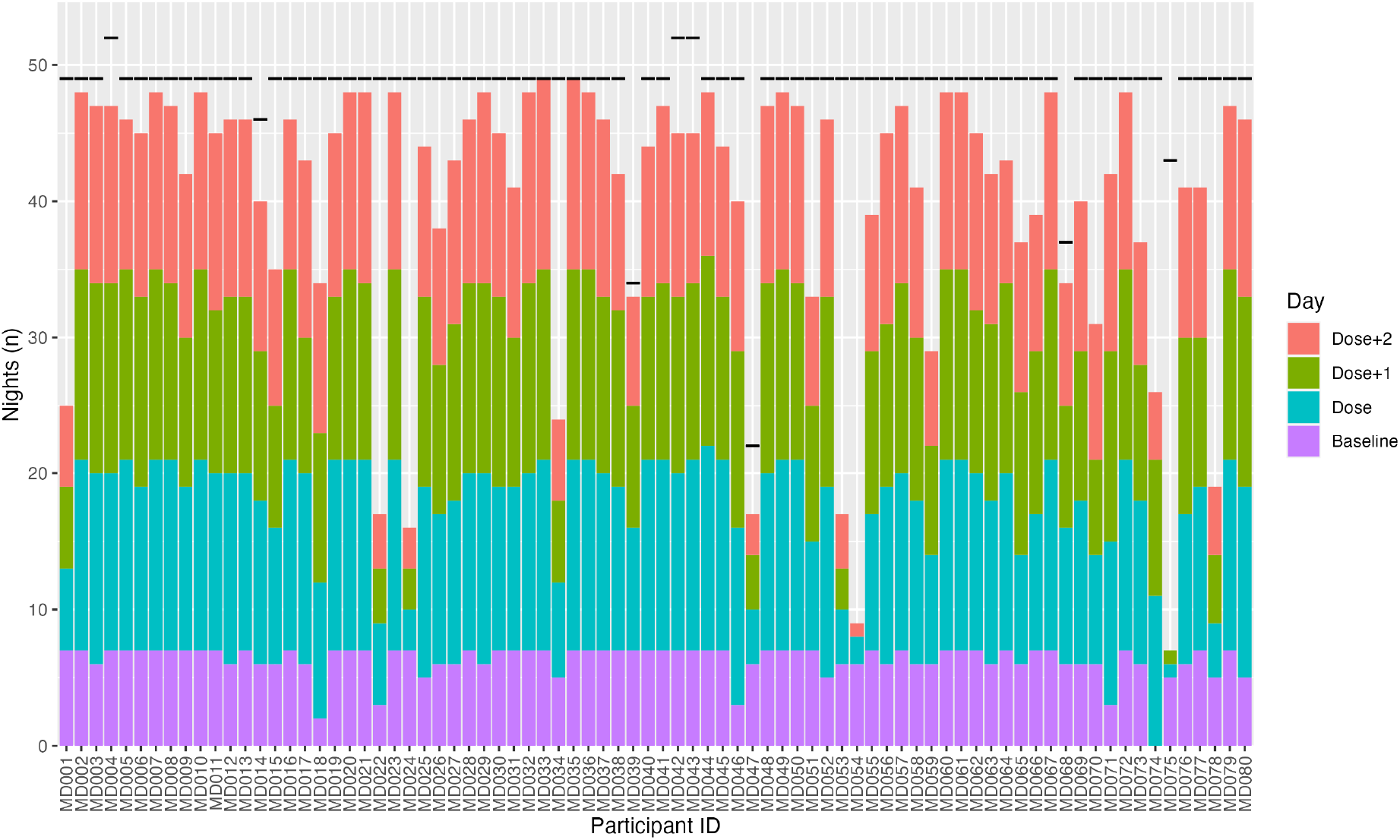
The number of days with quantifiable sleep data for each participant (MD001-MD080) characterised by baseline, dose day, dose+1 day, dose+2 day. See text for details.

**Supplementary Figure 2:**
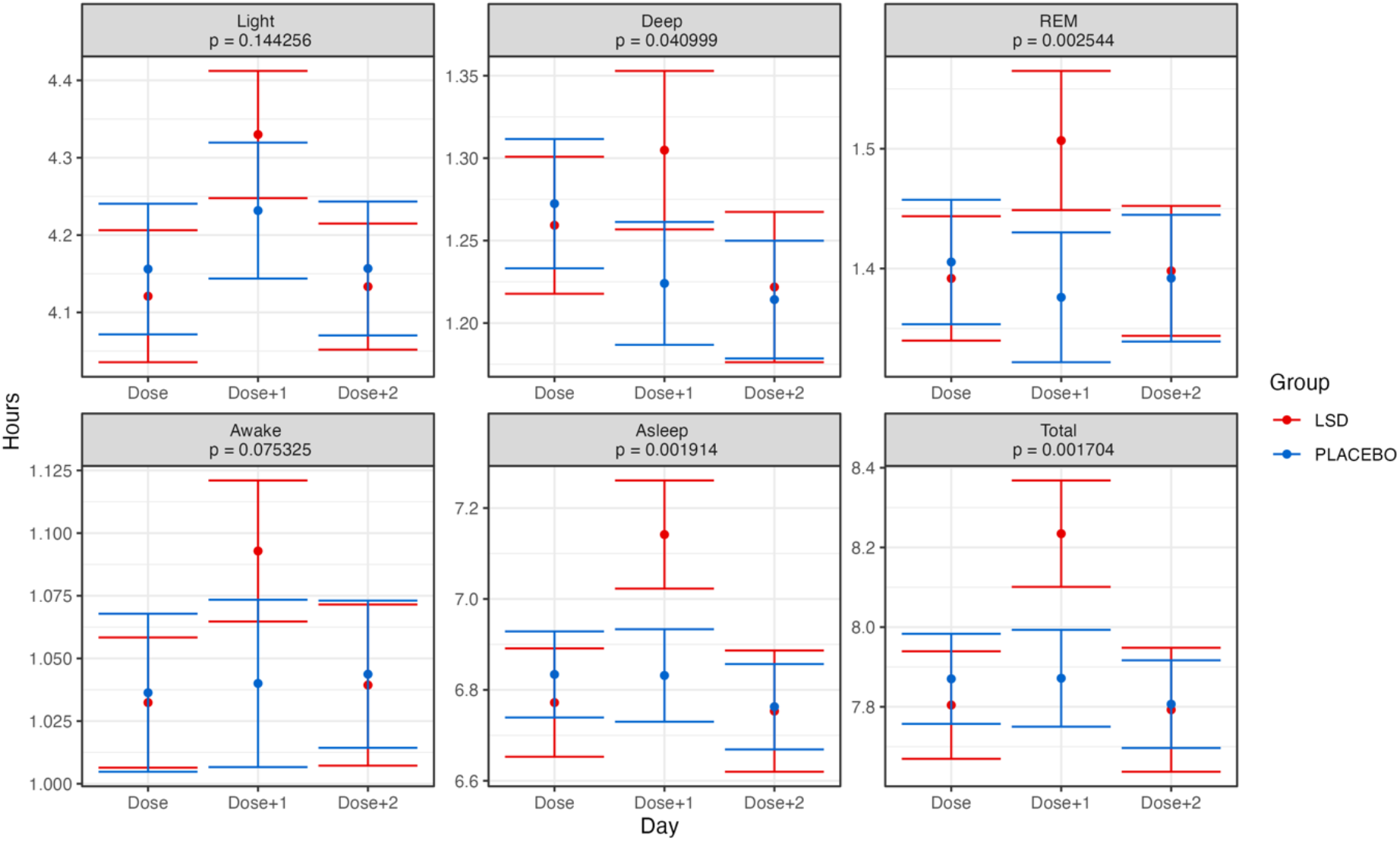
Grand average time spent in each of the sleep stages for each day (dose, dose + 1 dose + 2) and group in the trial. Error bars represent the standard error of the mean calculated across participants. Provided p-values are without baseline adjustment for reference.

**Supplementary Figure 3:**
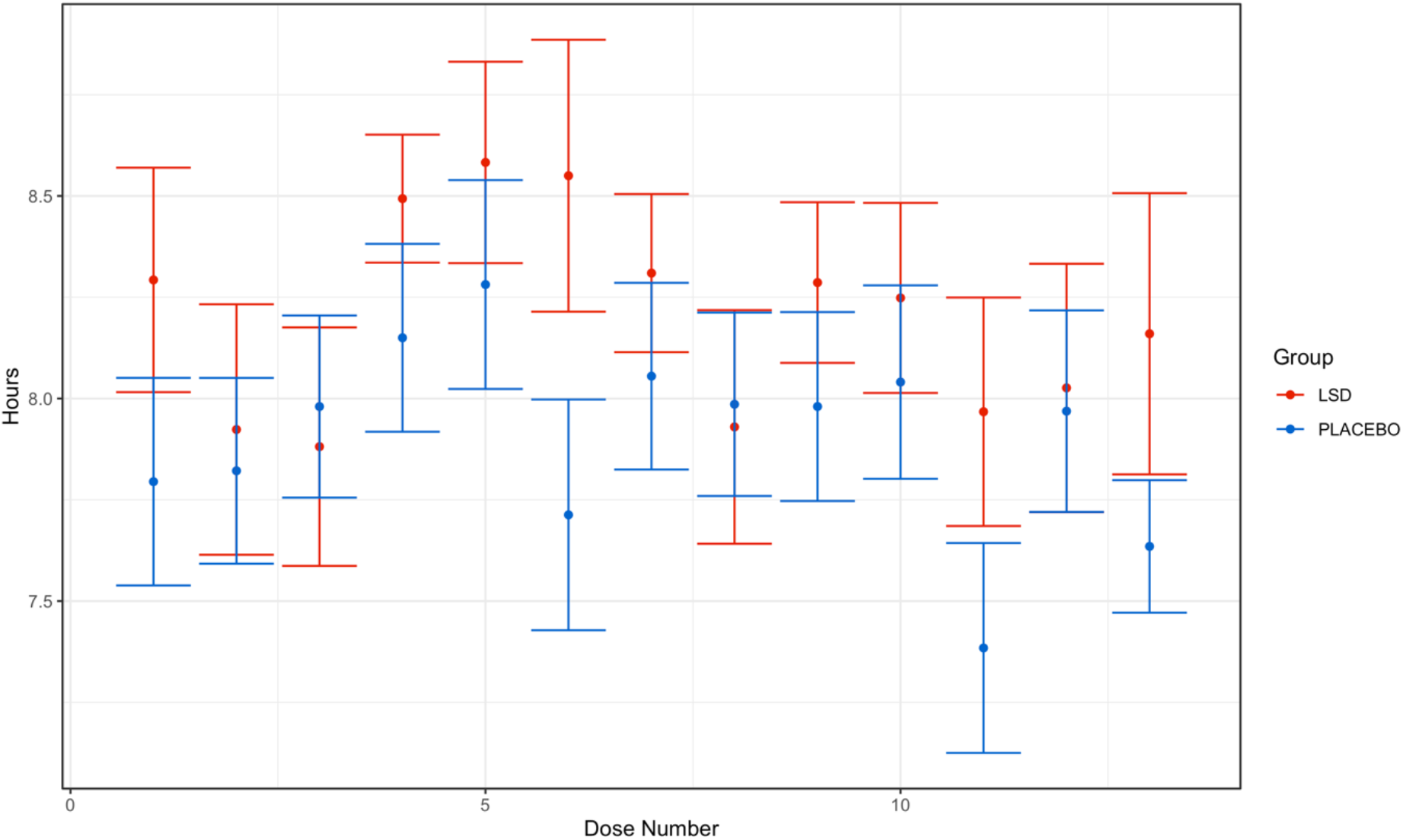
Grand average time spent in total sleep for each dose and group in the trial for the dose + 1 night. Error bars represent the standard error of the mean calculated across participants.

**Supplementary Figure 4:**
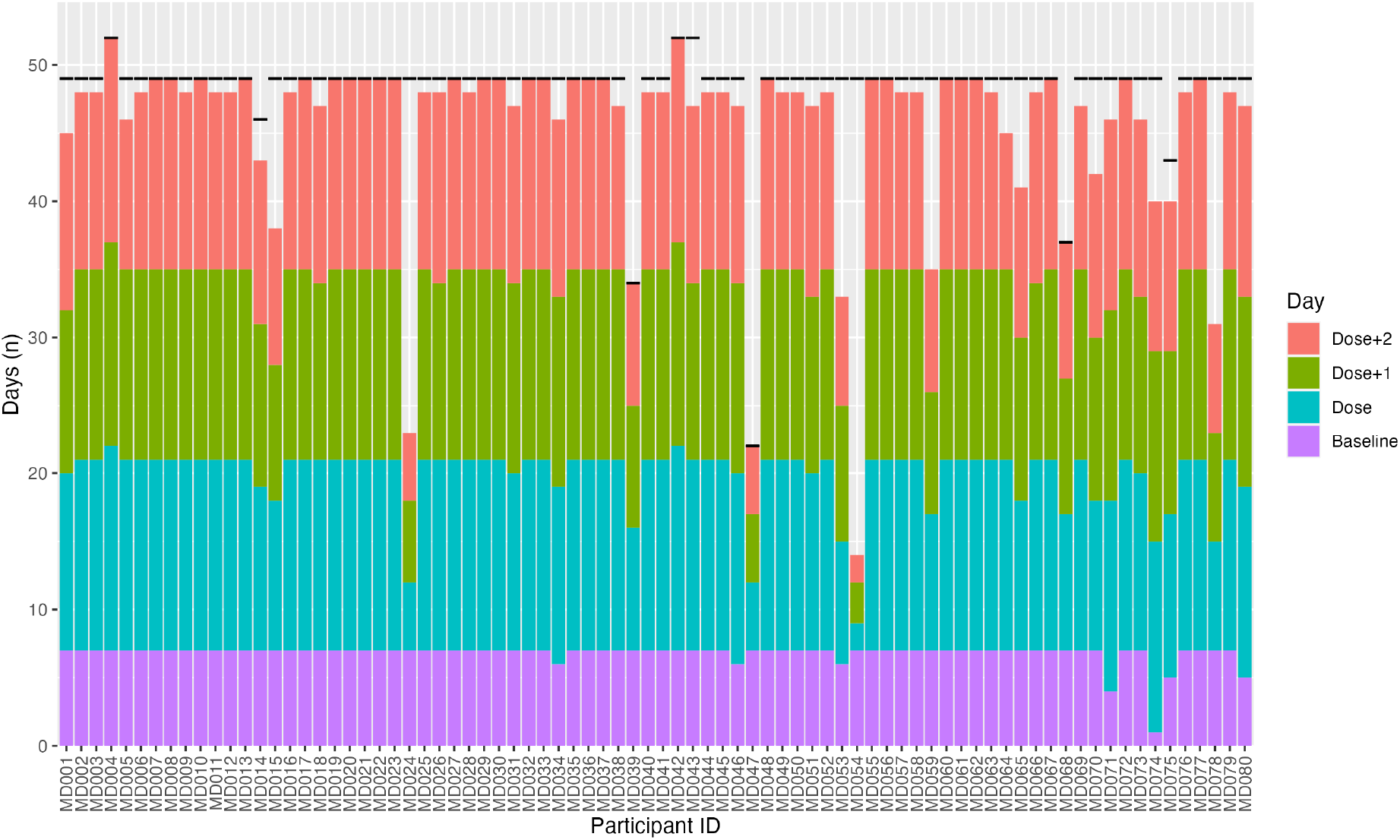
The number of days with quantifiable step data for each participant (MD001-MD080) characterised by baseline, dose day, dose+1 day, dose+2 day.

